# Left bundle branch pacing with and without anodal capture: impact on ventricular activation pattern and acute hemodynamics

**DOI:** 10.1101/2023.02.06.23285558

**Authors:** Nadine Ali, Khulat Saqi, Ahran D. Arnold, Alejandra A.Miyazawa, Daniel Keene, Ji-Jian Chow, Ian Little, Nicholas S. Peters, Prapa Kanagaratnam, Norman Qureshi, Fu Siong Ng, Nick W. F Linton, David C. Lefroy, Darrel P. Francis, PB Lim, Mark A. Tanner, Amal Muthumala, Matthew J. Shun-Shin, Graham D. Cole, Zachary I. Whinnett

## Abstract

**Introduction:** Left bundle branch pacing (LBBP) can deliver physiological left ventricular activation, but typically at the cost of delayed right ventricular (RV) activation. A proposed solution is to advance RV activation through anodal capture, but there is uncertainty regarding the mechanism by which early RV activation is achieved (capture of right bundle or RV myocardial capture) and it is not known whether this produces hemodynamic benefit.

**Methods:** We recruited patients with LBBP leads in whom anodal capture eliminated the terminal R wave in lead V1.

Ventricular activation pattern, timing and high precision acute hemodynamic response were studied during LBBP with and without anodal capture.

**Results:** We recruited 21 patients, mean age 67 years, 14 were males. We measured ECG timings and hemodynamics in all patients and in 15 we also performed non-invasive mapping. Ventricular epicardial propagation maps demonstrated that right ventricular septal myocardial capture, rather than right bundle capture, was the mechanism for earlier RV activation. With anodal capture, QRS duration was shorter (116 ± 12ms versus 129 ± 14ms, p < 0.01), and total ventricular activation time was shorter (83 ± 18ms versus 90 ± 15ms, p = 0.01). This required higher outputs (3.6 ± 1.9V versus 0.6 ± 0.2V, p <0.01) but did not provide additional hemodynamic benefit (mean difference −0.2 ± 3.8 mmHg compared to pacing without anodal capture, p = 0.2).

**Conclusion:** Left bundle pacing with anodal capture advances right ventricular activation as a result of stimulation of the RV septal myocardium. However, this requires higher outputs and did not improve acute hemodynamics. Aiming for anodal capture may therefore not be necessary.

## Introduction

Left bundle branch pacing (LBBP) may transform pacing therapy for both bradycardia and cardiac resynchronization^1–6^. Like His bundle pacing (HBP), it attains physiological left ventricular activation but offers several technical advantages:

- Pacing thresholds are lower and more stable ^3,7,8^
- Operators learn more quickly ^9^
- More distal conduction abnormalities can be treated ^7,8,10^

HBP is still considered the most physiological ventricular pacing modality ^11^. This is because HBP can produce physiological or near physiological biventricular activation^12^<u>. During LBBP,</u> right ventricular activation is typically delayed compared to HBP or normal intrinsic activation, this is manifest on the 12 lead ECG by a terminal R-wave (R-prime) in lead V1. The impact of this non-physiological right physiological activation on cardiac function and long-term outcomes is unknown. The observation of adverse outcomes in patients with right bundle branch block led to concerns that non-physiological right ventricular activation during LBBP could adversely impact cardiac function ^13,14^.

There are two proposed methods to overcome this delayed RV activation.

First, AV delay can be lengthened to allow intrinsic right bundle conduction to fuse with left bundle area capture ^4^. However, such fusion is not always achievable (e.g. in patients who have right bundle branch block, or complete heart block) or desirable ^15^ (e.g. if it requires a very long AV delay that impairs ventricular filling).

Second, the pacing output in bipolar configuration can be increased so that the anode stimulates the right ventricle. There is uncertainty regarding the mechanism through which anodal capture results in early RV activation. Some investigators have suggested this occurs as a result of capture of the right bundle branch (bi-bundle capture when combined with LBBP) whereas others propose it occurs through right ventricular myocardial capture ^4,16,17^. Crucially, it is not known whether programming the pacing output to obtain anodal capture, offers additional benefit with respect to hemodynamic function.

We investigated the mechanism of RV stimulation during LBBP with anodal capture, and compared the electrical and acute hemodynamic effects of LBBP with and without anodal capture.

## Methods

### Study design and patient recruitment

We enrolled consecutive patients with a left bundle branch pacing lead in whom it was possible to the eliminate R-prime in lead V1 with any pacing output during bipolar pacing. This involved bipolar pacing at sufficient outputs to capture the anode. To establish that the paced complexes were due to fusion of anodal and left bundle capture, we ensured that the ECG morphology was different compared to the pure anodal capture pattern of pacing between ring and pacemaker can.

### Left bundle pacing procedure

The transseptal approach described by Huang et al was used to implant the LBBP lead.^18^ We used the following criteria to confirm left bundle branch capture: (1) R prime in V1 during unipolar pacing, (2) Change in paced QRS morphology with changing pacing output or programmed stimulation during unipolar pacing, (3) Capture to R wave peak time in lead V5/V6 <90 ms.^19^

### Electrical measurements

#### 12 lead ECG

We used the Bard electrophysiology system (Boston Scientific, Natick, Massachusetts) to measure ECG parameters including QRS duration and R wave peak time. Threshold testing was carried out in VVI mode, during unipolar and bipolar configurations. The anodal capture threshold (loss of V1 R-prime) during bipolar pacing was noted.

In every patient the paced morphology at different AV delays was also examined, during pacing at the lowest output demonstrating left bundle capture. We adjusted the AV delay in 40ms increments, with the aim of identifying whether fusion with intrinsic conduction via the right bundle could be achieved (evidenced by loss of R-prime in V1).

#### Multi-electrode mapping

In some patients, selected opportunistically, multielectrode mapping was used to examine ventricular activation pattern, and measure ventricular activation times.

These patients wore a vest with 252 electrodes (ECGi, CardioInsight, Medtronic, Minneapolis, Minnesota).

Electrode location relative to each patient’s unique anatomy was accomplished using a low-dose thoracic computed tomography. The analysis was carried out using custom software in Python version 3.6 (Python Software Foundation, Wilmington, Delaware). For every parameter, the average of 5 beats each was taken.

Ventricular activation patterns, total ventricular activation time and left ventricular activation time were recorded and analyzed during the following;

1. Left bundle area only capture (R-prime present in V1); unipolar or bipolar pacing at the lowest output that achieved left bundle area capture.
2. Fusion between left bundle area capture and intrinsic conduction via the right bundle; pacing at the lowest output for left bundle area capture and the shortest AV delay resulting in fusion.
3. Left bundle area capture fused with anodal capture (no R prime in lead V1): bipolar configuration at the lowest output demonstrating fusion with anodal capture.

### Hemodynamic response

Hemodynamic response was assessed during LBBP with and without anodal capture using a high precision hemodynamic protocol ^20 21^. In brief this consisted of using beat-by-beat blood pressure measured either invasively or non-invasively. Each tested setting was compared to a reference setting. At least 6 alternations were made between the tested and reference setting. The analysis was automated and the mean change in SBP was calculated for each tested pacing configuration, relative to the reference setting, which was kept constant in an individual patient. The average of 8 beats was calculated at each alternation. To avoid any interruption to pacing we programmed a heart rate that was 10-15% above intrinsic rate.

### Ethics

All patients provided written informed consent. The study was approved by the health research authority (REC 19/YH/0174), the study was registered on ClinicalTrials.gov (NCT04221763).

### Statistical analysis

Continuous variables are expressed as mean and standard deviation or median and interquartile range. Categorical variables are expressed as proportions. For normally distributed variables a Student t-test was used for comparison, a paired Student t-test was used for dependent variables. P-values of <0.05 was considered statistically significant. Statistical analysis was conducted in RStudio using the tidyverse package.

## Results

### Patient population

We recruited 21 patients with mean age 67 years, 14 were male. The pacing indication was for bradycardia in 5 and cardiac resynchronization 16 patients. Demographics are shown in Table 1.

**Table 1.** demographics. Patient characteristics

| Table 1. Patient characteristics |  |
| --- | --- |
| **Demographics** |  |
| **Male** | 14 (67%) |
| **Age, y** | 67 ± 11 |
| **Device type** |  |
| **VVI** | 1 (5%) |
| **DDD** | 4 (19%) |
| **CRT-P** | 1 (5%) |
| **CRT-D** | 15 (71%) |
| **ECG Morphology** |  |
| **Normal** | 2 (10%) |
| **RBBB** | 3 (14%) |
| **LBBB** | 14 (67%) |
| **IVCD** | 1 (5%) |
| **Paced** | 1 (5%) |
| **ECG QRS duration / ms** | 157 ± 28 |
| **Cardiomyopathy** | 17 (81%) |
| **Ischemic** | 2 (12%) |
| **Non ischemic** | 15 (88%) |
| **LVEF %** | 36 ± 10 |

### Mechanism of loss of R prime evaluated using multielectrode epicardial propagation maps

We acquired multielectrode epicardial propagation maps in 15 of the 21 patients. This allowed us to assess the ventricular activation pattern during the two ventricular pacing configurations. During all forms of LBBP, left ventricular activation was rapid and physiological with the main ventricular activation wavefront proceeding from apex to base in 15/15 patients (which is consistent with activation via the left conduction system). The pattern of RV activation differed between the two forms of LBBP (Figures 1,2 and 3). During LBBP without anodal capture, RV activation occurred via two wavefronts, one which was most likely due to septal breakthrough from local left ventricular myocardial septal capture (from the lead tip) and the second occurred via a posterior wavefront (likely breakthrough from the LV conduction system capture). The basal free wall of the RV was the latest activated area in all patients. 15/15 (Figure 1).

**Figure 1.**
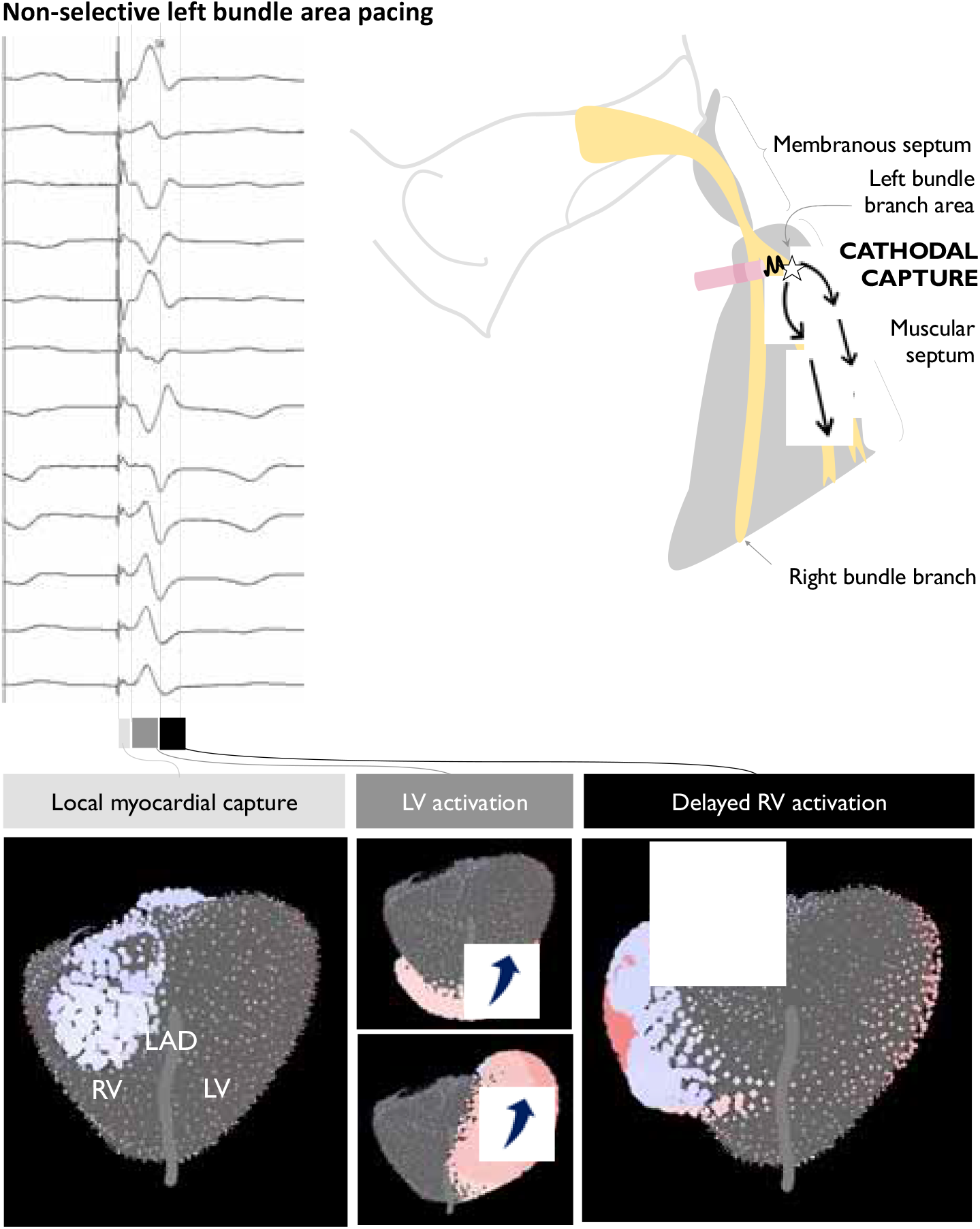
Activation during non-selective left bundle branch capture. Activation starts with local myocardial capture. The left ventricle activates in a physiological fashion from apex to base. There is delayed activation of the right ventricle with a region of the basal free wall activating late and this is represented by the terminal R wave in lead V1.

**Figure 2.**
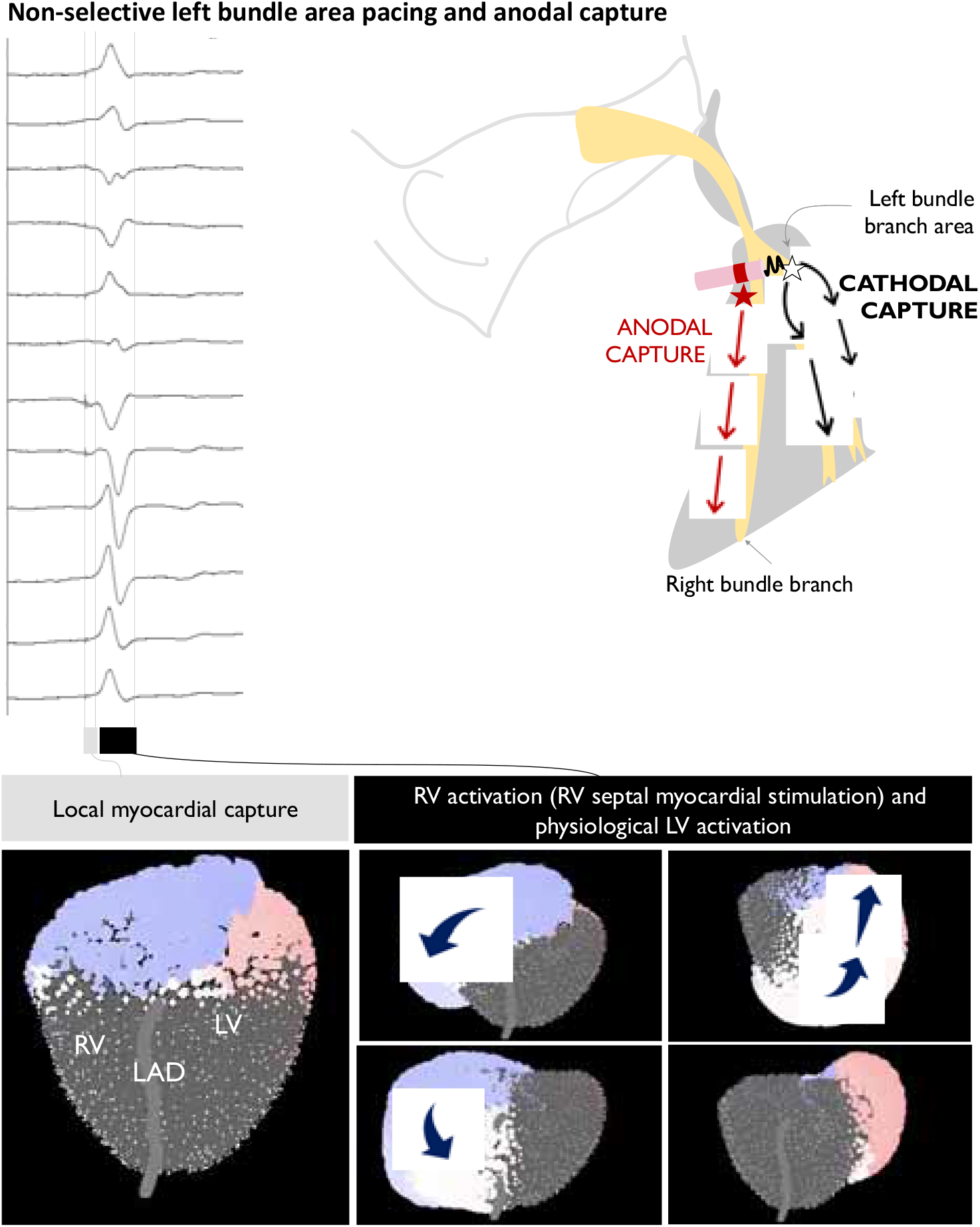
Non-selective left bundle branch capture. A large area of the RV septum activates first with activation spreading from the septum, right ventriclar activation occurs from base to apex (i.e. different to intrinsic activation via the right bundle). We typically also observed RV acivation via a posterior wavefront (likley breakthrough from LV activation via the left conduction system). Acitvation of the basal lateral wall of the right ventricle is advanced compared to LBBP without anodal capture. The left ventricle activates in a physiological fashion from apex to base.

**Figure 3.**
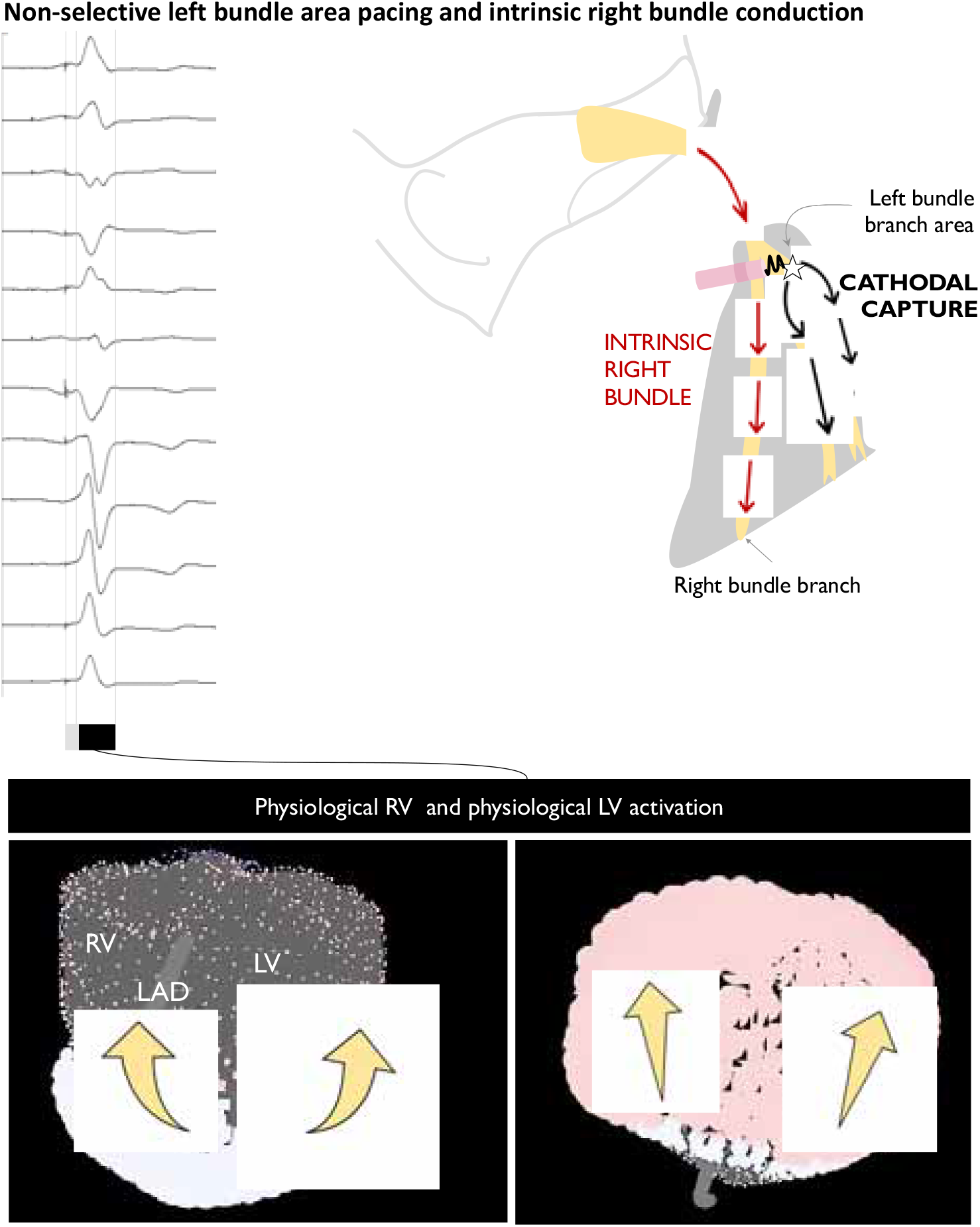
Non-selective left bundle branch capture fused with intrinsic right bundle conduction. The right and left ventricles activate in a physiological fashion from apex to base.

LBBP with anodal capture, resulted in advancement of RV activation compared to activation without anodal capture in 15/15. The earliest area of right ventricular activation was basal RV (due to RV septal myocardial activation), with activation spreading from base to apex of the RV i.e. not consistent with activation via the right bundle. A second posterior wavefront was also observed (likely due to breakthrough from left conduction system activation). The activation of the basal lateral wall was advanced compared to pacing without anodal capture, which appeared to occur because of earlier RV septal activation.

Therefore, the propagation maps suggested that that anodal capture advanced right ventricular activation via septal myocardial capture rather than bi-bundle capture, in all of our patients. (Figure 2).

During LBBP fused with intrinsic right bundle activation (achievable in 6/15), RV activation was rapid and consistent with physiological activation via the right bundle, with the activation wavefront propagating from RV apex to base (Figure 3).

### Comparison of Ventricular activation times: LBBP with and without anodal capture

LBBP with anodal capture produced a modest reduction in 12-lead ECG QRS duration (−12 ± 7.0 ms) compared to LBBP without anodal capture (mean QRS duration 129 ± 14ms without anodal capture and 116 ± 12ms with anodal capture, p < 0.01, Figure 4).

**Figure 4.**
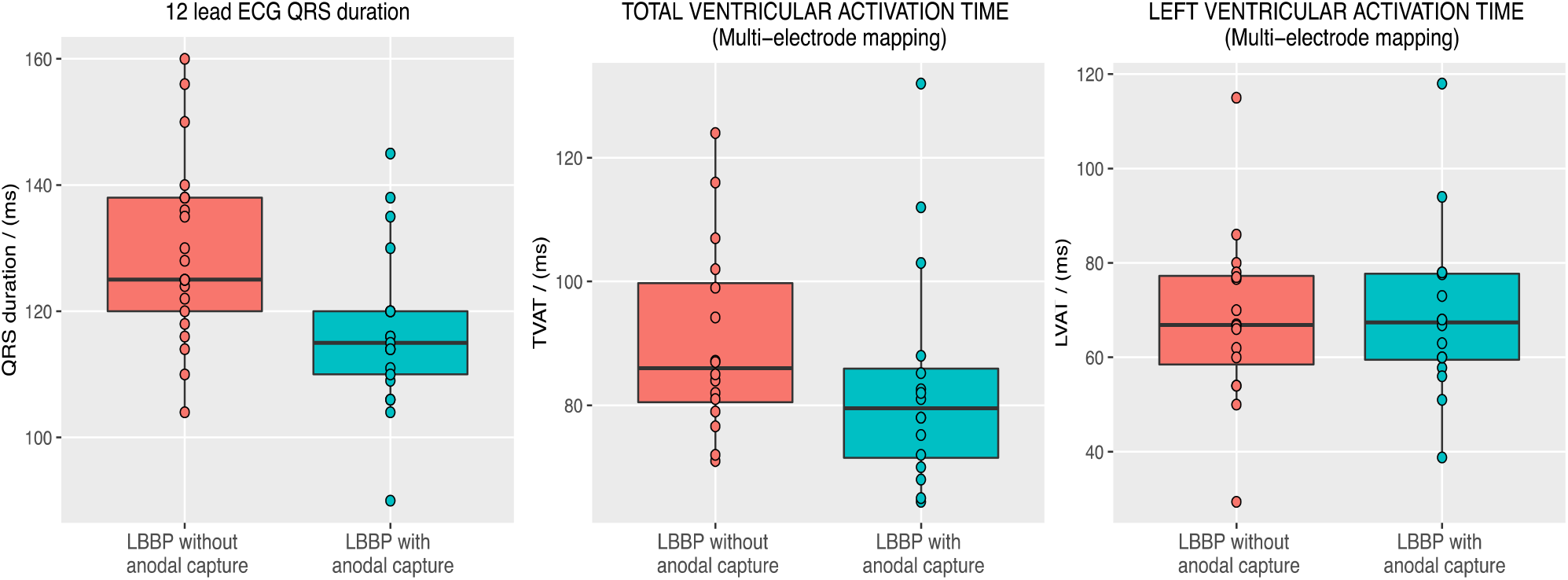
**Left;** QRS duration (12 lead ECG) produced by Left bundle branch pacing without and with anodal capture, there was a modest reduction in QRS duration associated with anodal capture. **Middle;** Total ventricular activation time (TVAT) (measured with multi-electrode mapping) with and without anodal capture, there was a modest reduction in TVAT with anodal capture. **Right;** Left ventricular activation time (LVAT) (measured with multi-electrode mapping) with and without anodal capture, there was no difference in LVAT between the two configurations.

In the patients in whom multi-electrode mapping was available (15/21) we observed a modest reduction in total ventricular activation time (TVAT) of (−7 ± 9ms) with anodal capture compared to LBBP without anodal capture (TVAT 90 ± 15ms without anodal capture and 83 ± 18ms with anodal capture, p = 0.01, Figure 4). Left ventricular activation times did not differ between the two pacing configurations (68 ± 19ms and 69 ± 18ms, p = 0.6, Figure 4). LBBP fused with intrinsic right bundle conduction was observed in 6/15 patients, multi-electrode mapping was available in all 6. We observed a modest reduction in TVAT (12 ± 6ms) during LBBP fused with intrinsic right bundle conduction, compared to LBBP without fusion (TVAT 78 ± 13ms Vs TVAT 90 ± 16ms, p < 0.05, Figure 5). Left ventricular activation times did not differ between the two pacing configurations (62 ± 3ms and 59 ± 8ms, p = 0.4, Figure 5).

**Figure 5.**
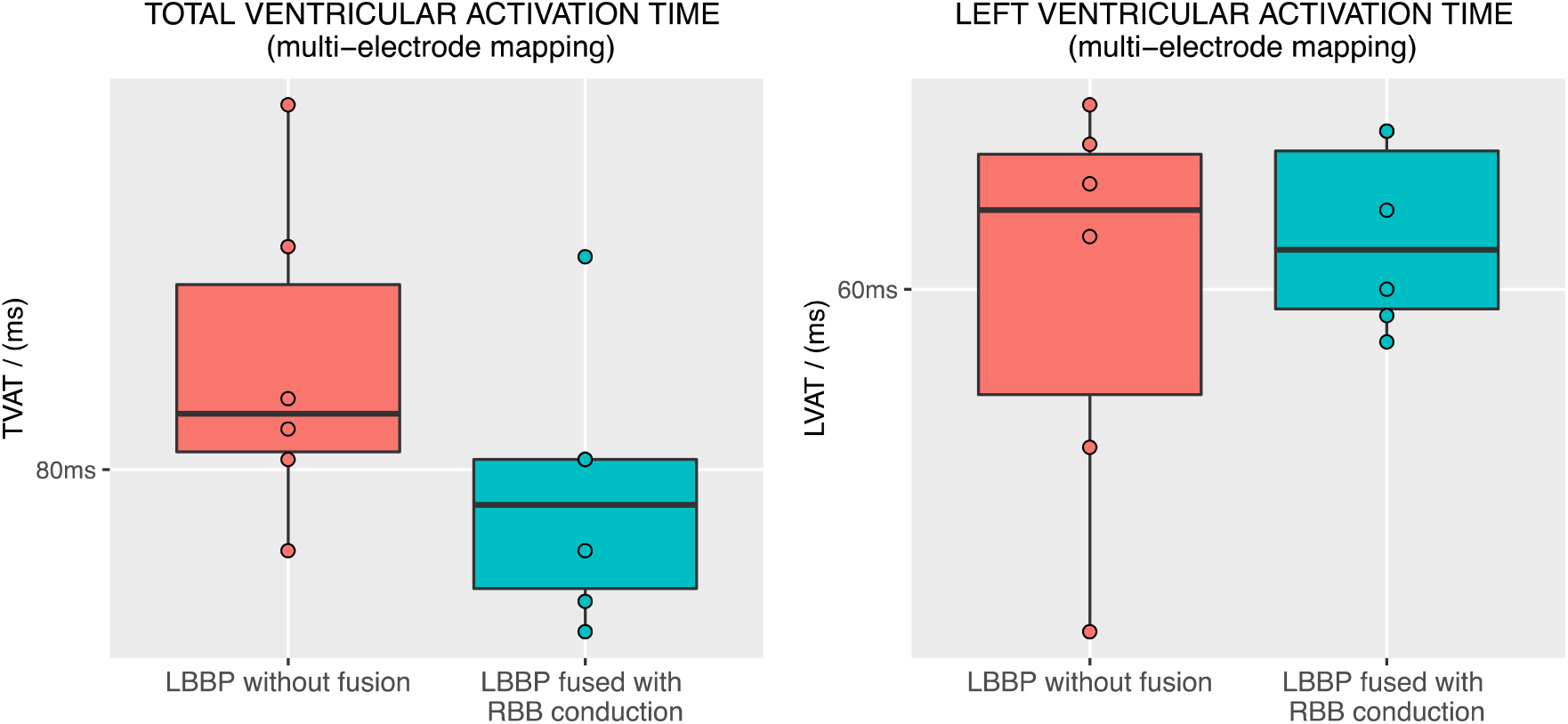
**Left;** Total ventricular activation time (TVAT) (measured with multi-electrode mapping) with and without fusion with intrinsic right bundle conduction, there was a modest reduction in TVAT with fusion with right bundle branch conduction. **Right;** Left ventricular activation time (LVAT) (measured with multi-electrode mapping) with and without fusion with intrinsic right bundle conduction, there was no difference in LVAT between the two configurations.

### Hemodynamic response

High precision hemodynamic response was assessed in all 21 patients. There was no significant hemodynamic difference between LBBP capture only compared to LBBP with anodal capture: −0.2 ± 3.8 mmHg, p = 0.2 (Figure 6).

**Figure 6.**
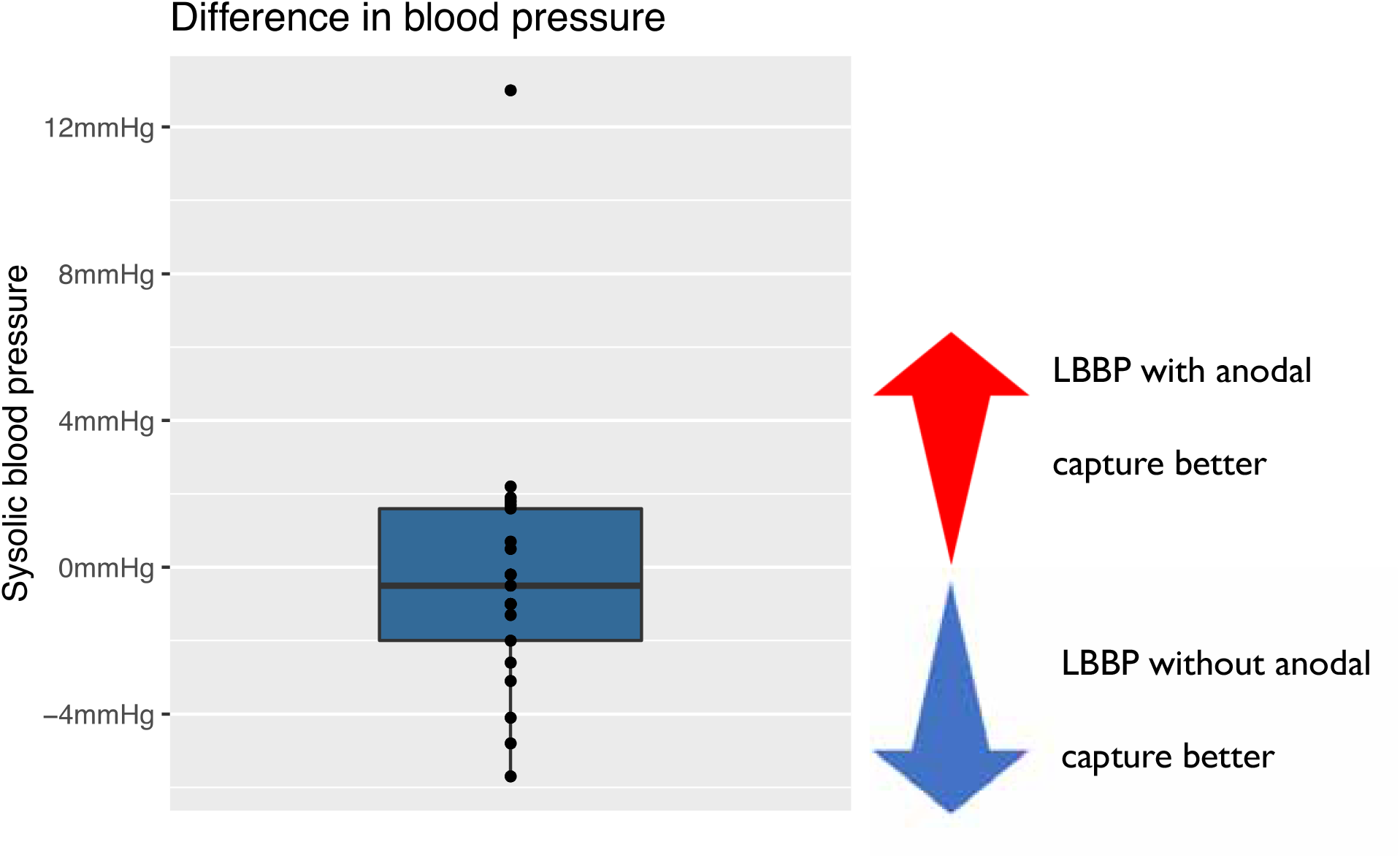
Hemodynamic comparison of LBBP with and without anodal capture. We found no difference in acute hemodynamic response during left bundle pacing with and without anodal capture.

### Pacing thresholds

LBBP with anodal capture required higher pacing outputs, mean capture threshold was 3.6 ± 1.9V at 0.9 ± 0.2ms compared to 0.6 ± 0.2V at 0.8±0.3ms for LBBP without anodal capture (p <0.01, Figure 6).

**Figure 6.**
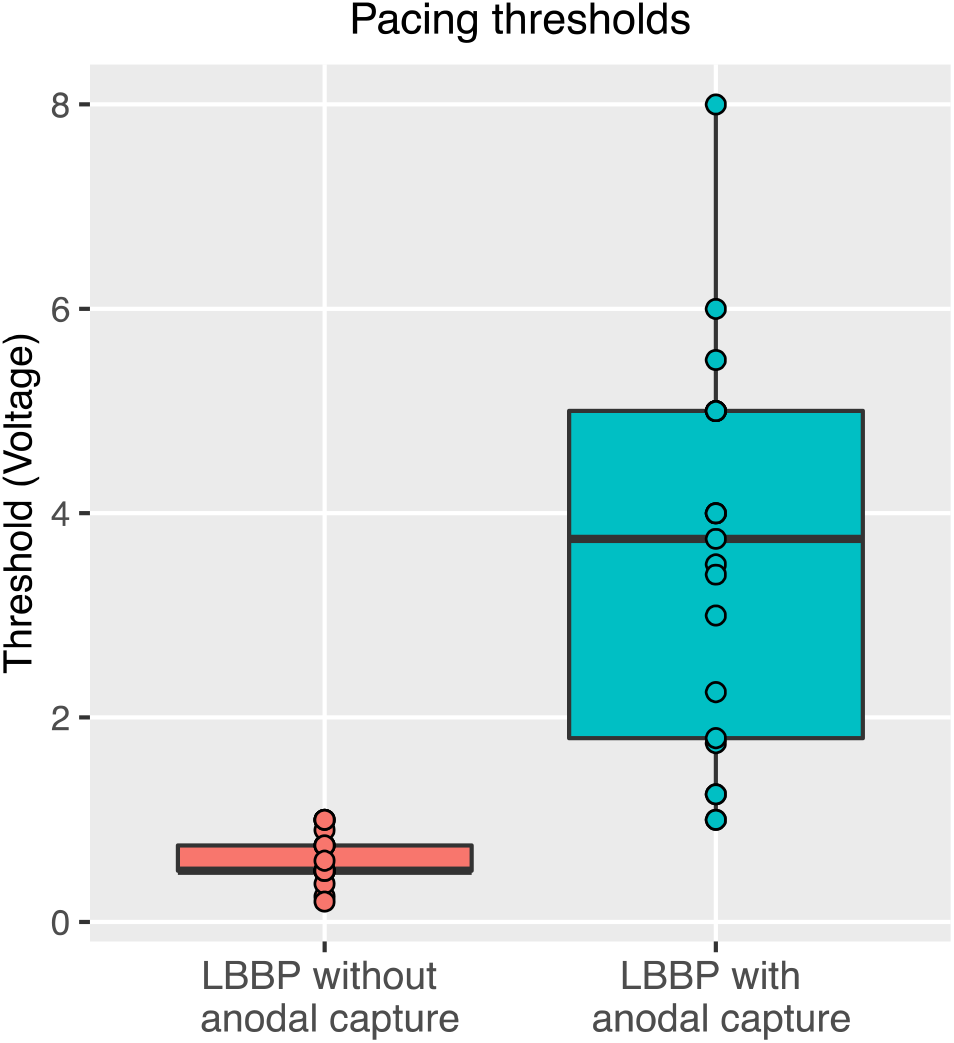
Pacing threshold for left bundle branch pacing with and without anodal capture. Achieving anodal capture in addition to left bundle area capture required significantly higher pacing outputs.

## Discussion

This study confirms that achieving anodal capture during LBBP produces modest improvements in ventricular activation times, compared to LBBP without anodal capture. Our findings suggest that activation time was reduced by advancing right ventricular myocardial septal capture, rather than direct capture of the right bundle (bi-bundle capture) in all of our patients in whom we collected ventricular epicardial propagation maps. Achieving anodal capture required higher pacing outputs and even with high precision measurements, there was not even the slightest sign of better acute hemodynamics. These findings suggest that adding anodal capture improves the electrical appearance without any immediate benefit in cardiac performance.

### Mechanism of early right ventricular activation

Our study is the first to apply multi-electrode mapping in order to address the question of whether anodal capture during LBBP advances RV activation by capturing the right bundle branch or through right ventricular septal myocardial capture.

In all 15 patients, in whom we performed multielectrode mapping the activation pattern was consistent with right ventricular septal capture, rather than right bundle capture.

The reason for this may be that the left and right bundles do not pass down the septum in mirror image positions, but rather the right bundle typically courses more anteriorly and superiorly than the left bundle. Therefore, when the lead is passed through the septum to the left bundle area, its trajectory does not generally pass through the right bundle. With the tip of the lead positioned an average of 14mm into the septum ^22^, and the ring (anode) of the commonly used SelectSecure lead being 9mm from the tip, the anodal stimulation site is typically around 5mm into the septum. This position is generally well within the right ventricular septal endocardium and too posterior to capture the right bundle directly.

### Hemodynamic response

Disappointingly, despite the clear advancement of right ventricular activation, narrowing of the QRS and shortening of total activation time, there was no evidence of any hemodynamic advantage. Indeed, the numerical values were a non-significant trend towards worse hemodynamics with anodal capture. The high precision hemodynamic protocol we used can detect even small hemodynamic advantages through the use of large numbers of repeated transitions, automated beat alignment and standardized statistics. Therefore, it seems very unlikely that adding anodal capture to LBBP will ever provide improvements in acute hemodynamics.

We speculate that the reason for this lack of hemodynamic benefit is that while anodal capture does advance RV activation compared to LBBP without anodal capture, this only produces a modest reduction in activation time by advancing the latest area of activation (RV basal septum). Overall, right ventricular activation is similar between the two types of LBBP capture, for both capture types: 1) activation spreads via myocardial activation rather than utilizing the conduction system 2) RV activation is produced by two wavefronts, one originating from the septum (this is delayed in the absence of anodal capture) and the second due to posterior breakthrough from the left ventricular conduction system-initiated activation. The main difference between the two capture types is a small area of late activation in the basal free wall which is observed in the absence of anodal capture. The findings of our study suggest that this small area of late activation has no impact on acute hemodynamic function.

### Implications for device programming

During BVP pacing physicians often aim to program the device to achieve the narrowest possible QRS. However, the findings of our study suggest that for LBBP, while adding anodal capture shortens QRS duration it does not improve acute hemodynamic function. This narrower QRS often comes at a substantial cost in terms of pacing output required and therefore battery life.

Therefore, our findings suggest that outside of adequately powered randomized controlled trials of long-term effects, we should not routinely program anodal capture for patients in whom this requires many-fold greater pacing output. This is the great majority of patients.

### Limitations

The multi-electrode mapping equipment was only available in 15/21 patients, due to funding limitations. However, the activation patterns seen were consistent in all 15 patients. The number of patients recruited was appropriate for a physiological study and delivered narrow confidence intervals as planned. However, this is small compared to the sample size required for an event study where each patient contributes only one binary digit of information.

This was an acute study, we did not investigate the chronic impact of LBBP with and without anodal capture. Whether the addition of anodal capture results in any long-term benefits would need to be investigated in a separate study.

The multielectrode mapping technique, ECGi, can only report epicardial activation and not endocardial. Invasive mapping would be required to address this. Nevertheless, we believe the information from the ECGi is persuasive that the activation is through septal myocardial stimulation, rather activation via the right bundle (in which case we would to see apex to base RV activation as we did during intrinsic activation).

In this study, we used long programmed AV delays as one method of achieving fusion between paced left ventricular activation and native right ventricular activation. However, this was purely an experimental method of temporarily achieving fusion, and not a recommendation to program this chronically in routine practice. Programming long AV delays in patients with a long intrinsic PR interval may adversely affect ventricular filling which may offset the beneficial effects of intrinsic right ventricular activation ^15^.

In this study we did not perform a direct comparison of intrinsic conduction via the right bundle with the activation achieved via LBBP, which should be performed at the same AV delay, for example with HBP. We cannot therefore answer the question of whether RV activation via the intrinsic conduction system produces superior acute hemodynamics to LBBP pacing initiated RV activation. The aim of our study was to establish the impact of anodal capture.

## Conclusions

Adding anodal capture to LBBP does attenuate the delay in right ventricular activation, and the mechanism is stimulation of the right ventricular septal myocardium rather than capture of the right bundle branch. There is clear but modest shortening of QRS duration and total ventricular activation time. However, this requires an approximately six-fold higher pacing output and shows not even the slightest sign of improving acute hemodynamics. Our findings do not support routinely programming anodal capture during LBBP.

## Data Availability

Data is available upon reasonable request.

